# Caregiver-Rated Inappropriate Speech and Post-cTBS Motor Cortical Facilitation in Autism: A Pilot Biomarker Study

**DOI:** 10.64898/2026.09.10.26362749

**Authors:** Joshua Ryan Smith, Maria Bonnee, Sarah Marler, Rowan Atwood, Brianna Lewis, Seri Lim, Isaac Baldwin, Hao Wu, Jinyuan Liu, Carissa Cascio, Gagan Joshi, Paul E. Croarkin

## Abstract

Transcranial magnetic stimulation can provide noninvasive measures of cortical excitability and plasticity, but relationships between these measures and clinically observable features of autism are not fully characterized. Nineteen autistic participants aged 15 to 40 years were included in the analysis of a left primary motor cortex continuous theta-burst stimulation (cTBS) biomarker protocol. Motor evoked potentials were measured at baseline and at seven assessments from 5 to 60 min after stimulation. Linear mixed-effects models evaluated whether clinical measures moderated the post-stimulation motor evoked potential log-response ratio over time. Aberrant Behavior Checklist (ABC) and Attenuated Behavior Questionnaire (ABQ) analyses were restricted to the same 15 participants with caregiver-informant assessments. Catatonia severity, social impairment, ABQ Motor Total, ABQ Total, and cognitive ability did not significantly moderate the post-stimulation response. ABC Inappropriate Speech was associated with progressively greater post-stimulation facilitation (standardized Time × Inappropriate Speech interaction: β = +0.688, SE = 0.210, 95% CI +0.276 to +1.099; Holm-adjusted P = .008 across eight informant-rated models). Two individual speech-related items (“Talks excessively” and “Talks to self loudly”) survived false-discovery-rate correction. An exploratory eight-item ABC phenotype showed a large descriptive in-sample association (β = +0.904, SE = 0.200, P < .001) and directionally positive held-out performance. However, full-pipeline permutation tests were nonsignificant for both Pearson (r = .357, two-sided empirical P = .360) and Spearman correlations (ρ = .446, two-sided empirical P = .261). Caregiver-rated inappropriate speech may be associated with altered post-cTBS motor cortical facilitation in autism. The candidate phenotype remains hypothesis-generating and requires independent validation.

## Introduction

Autism is a heterogeneous neurodevelopmental condition associated with substantial psychiatric, medical, and functional morbidity [1–5]. Approximately 30–40% of autistic individuals have co-occurring intellectual disability [6–9], yet individuals with intellectual disability and high support needs remain markedly underrepresented in autism research [10,11]. This exclusion limits the generalizability of clinical and neurobiological findings and constrains biomarker development across the full autism spectrum.

There are no clinically established biomarkers to guide the diagnosis or treatment of autism or its associated conditions [12,13]. One prominent framework is excitation-inhibition (E/I) imbalance, in which altered inhibitory signaling may contribute to cortical hyperexcitability, atypical synaptic plasticity, and behavioral heterogeneity [13–16]. Transcranial magnetic stimulation (TMS) studies have reported altered cortical modulation, inhibition, and plasticity-like responses in autistic individuals [13,15,17–23], although many included predominantly participants without co-occurring intellectual disability.

TMS is a form of noninvasive brain stimulation with utility for discovery science focused on biomarker and intervention development. TMS has an established safety record in both adults and children [24–30]. When applied to the primary motor cortex (M1), TMS-evoked motor potentials quantify corticospinal excitability [13,31–35], while theta-burst stimulation (TBS) probes plasticity-like responses. M1 cTBS generally suppresses motor-evoked potential amplitude, although individual responses vary. Altered post-cTBS modulation in autism may therefore indicate atypical cortical plasticity [12,36].

Catatonia is increasingly recognized in autistic and neurodevelopmentally disabled individuals and may involve severe functional decline, mutism, posturing, aggression, and self-injury [37–47]. Proposed mechanisms include altered GABAergic and glutamatergic signaling and abnormal cortical excitability, partially overlapping with excitation-inhibition models of autism [16,48]. These shared mechanisms raise the possibility that catatonic and severe behavioral symptoms in autism may be associated with altered cortical plasticity.

Based on proposed abnormalities in inhibitory and excitatory signaling and cortical plasticity in autism and catatonia, we hypothesized that greater catatonia severity and greater social impairment would be associated with greater post-cTBS MEP facilitation, or less suppression, over time. Planned exploratory analyses evaluated attenuated catatonic behaviors, cognitive ability, and caregiver-rated aberrant behavior. Additional exploratory analyses characterized a candidate phenotype arising from the Aberrant Behavior Checklist (ABC) Inappropriate Speech finding [49] and evaluated its stability and selection-adjusted performance.

## Methods and Materials

### Study Design and Participants

This single-site, single-arm, nonrandomized Phase 1 diagnostic TMS biomarker study was conducted at Vanderbilt University Medical Center (VUMC) without a concurrent comparator. The VUMC Institutional Review Board approved the protocol (220645), and the study was registered on ClinicalTrials.gov (NCT06016764). Participants or legally authorized representatives provided written informed consent, with assent obtained when appropriate. The study followed the Declaration of Helsinki. Participants enrolled from August 2023 through July 2025.

Participants were 15 to 40 years old and met Diagnostic and Statistical Manual of Mental Disorders, Fifth Edition criteria for autism spectrum disorder [1], confirmed using the Autism Diagnostic Observation Schedule, Second Edition. Participants with and without catatonic features were recruited from Vanderbilt clinical programs.

Exclusion criteria were substance use disorder; major medical or neurological illness; seizure within one year; traumatic brain injury; pregnancy or breastfeeding; psychiatric or medical instability; TMS-contraindicated medications; prior TMS treatment; an autism-associated genetic syndrome expected to produce a distinct biomarker profile, including Fragile X syndrome; magnetic resonance imaging contraindications; or sustained dissent.

Twenty-six participants enrolled, 20 completed the diagnostic TMS procedure, and 19 were included in the primary analysis after exclusion of one participant assessed before implementation of the finalized tolerability-related protocol. Participant flow is shown in Supplementary Figure 1. No serious adverse events occurred. As a pilot biomarker study, the sample size was based on feasibility rather than a formal power calculation.

### Clinical and Behavioral Assessments

Catatonia severity was assessed using the Bush-Francis Catatonia Rating Scale (BFCRS) [50], Kanner Catatonia Rating Scale, represented by the Kanner Catatonia Severity (KCS) Total and Kanner Catatonia Examination (KCE) Total [51]. Social impairment was assessed using the Social Responsiveness Scale, Second Edition (SRS-2) Informant Total Raw Score [52]. Cognitive ability was assessed using the Wechsler Abbreviated Scale of Intelligence Second Edition (WASI-II) [53] Full Scale-4 IQ composite. For one participant, IQ scores were obtained from an assessment completed within 12 months before enrollment.

Attenuated catatonic behaviors were assessed using the caregiver-rated Attenuated Behavior Questionnaire (ABQ) [54]. The present analyses evaluated ABQ Motor Total and ABQ Total scores, with higher scores indicating greater attenuated catatonic symptom burden.

Aberrant behavior was assessed using the caregiver-rated ABC which yields Irritability, Social Withdrawal, Stereotypic Behavior, Hyperactivity/Noncompliance, and Inappropriate Speech subscale scores and an ABC Total Score. Items are rated from 0 to 3, with higher scores indicating greater symptom severity [49].

SRS-2 Informant scores were available for all 19 participants. Concurrent caregiver-rated ABC and ABQ data were available for 15 participants, who formed the multimeasure cohort. One participant was missing abc22 but had data for the other seven candidate-phenotype items.

### Transcranial Magnetic Stimulation Procedures

Stimulation was delivered using a figure-of-eight coil connected to a MagPro X100 stimulator (MagVenture, Farum, Denmark). The left primary motor cortex hand representation was first localized using frameless stereotactic neuronavigation registered to each participant’s structural magnetic resonance image. Within this anatomically defined region, coil position was then refined to identify the location producing maximal motor-evoked potentials in the right first dorsal interosseous muscle. This functionally defined location was used for subsequent stimulation.

Resting motor threshold was the minimum intensity producing an MEP of at least 50 μV on at least 5 of 10 trials at rest. Active motor threshold was the minimum intensity producing an MEP of at least 200 μV on at least 5 of 10 trials during contraction at 20% of maximal voluntary force. Baseline excitability was assessed using 30 single-pulse stimuli at 120% of resting motor threshold. cTBS was delivered over left M1 at 80% of active motor threshold as 600 pulses over 40 s. Ten single-pulse stimuli were administered at 5, 10, 20, 30, 40, 50, and 60 min after cTBS.

During study conduct, the TMS procedure was modified to improve procedural tolerability by reducing cTBS intensity to 80% of active motor threshold. Data from one participant collected before implementation of the finalized procedure were excluded from the analysis. The finalized procedure described above was used for all participants included in the analytic cohort.

### MEP Data Processing

Non-positive or nonnumeric motor evoked potential amplitudes were excluded. Within each assessment block, amplitudes exceeding 2.5 standard deviations from the block mean were removed, and the arithmetic mean of retained amplitudes was calculated.

The primary outcome was the post-cTBS motor evoked potential log-response ratio, denoted ΔMEP:

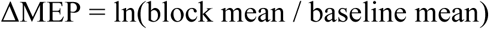

Positive values indicate facilitation relative to baseline, whereas negative values indicate suppression. This produced one ΔMEP value per participant at each of the seven post-cTBS assessments.

### Statistical Analysis

Analyses were performed in Python using pandas, NumPy, SciPy, statsmodels, and joblib. All tests were two-sided, with P < .05 defining statistical significance.

Separate linear mixed-effects models evaluated the association of each clinical predictor with the post-cTBS ΔMEP trajectory. Each model included all seven post-cTBS ΔMEP observations per participant, rather than a peak, average, or single selected time point. Time was modeled continuously and scaled from 0 at 5 min to 1 at 60 min. Participant-level predictors were standardized within the applicable analytic sample. Models included baseline MEP amplitude, time, the predictor, and the Time × Predictor interaction as fixed effects, with a participant-specific random intercept:

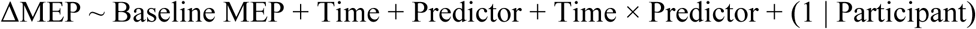

Models were estimated using restricted maximum likelihood. The Time × Predictor coefficient represents the difference in the ΔMEP trajectory from 5 to 60 min associated with a one-standard-deviation increase in the predictor. Positive coefficients indicate progressively greater facilitation, or less suppression, over the post-cTBS assessment period.

Prespecified Aim 1 separately evaluated BFCRS Total, KCS Total, and KCE Total in the full 19-participant cohort. Prespecified Aim 2 evaluated SRS-2 Informant Total Raw Score in the full cohort. Planned exploratory models evaluated WASI-II Full Scale-4 IQ in the full cohort and ABQ Motor Total, ABQ Total, the five ABC subscales, and ABC Total Score in the common 15-participant caregiver-informant cohort. Holm correction was applied across the eight caregiver-informant ABQ/ABC models. The prespecified clinical aims and directional hypotheses were unchanged; the ABC item-level, candidate-phenotype, nested-validation, multimeasure, permutation, and domain analyses were exploratory or post hoc. The analysis hierarchy, analytic samples, outcomes, multiplicity procedures, and inferential status are summarized in Supplementary Table 6.

Microsoft 365 Copilot assisted with workflow organization, Python code development and debugging, figure code, documentation, manuscript editing, and formatting. The authors independently reviewed and verified all analytic decisions, code, outputs, tables, figures, and text and take responsibility for the work.

### ABC Item-Level Analysis and Candidate Behavioral Phenotype

Following the ABC subscale analyses, exploratory models separately evaluated each of the 58 ABC items in the 15-participant caregiver-informant cohort. Each item was entered as the predictor in the same linear mixed-effects model described above, using all seven post-cTBS ΔMEP observations per participant. Benjamini-Hochberg correction was applied across the 58 Time × Item interaction tests. This procedure was an item-level screen, not a factor analysis or latent-variable model.

The eight items with the smallest raw Time × Item interaction *P* values were combined into an exploratory candidate phenotype spanning speech, motor or behavioral disinhibition, and self-injury content. Items were direction-aligned according to their observed Time × Item coefficients, standardized across participants, and averaged with equal weighting. Candidate-phenotype scores required at least six available items. All eight observed-sample coefficients were positive, so no items were reversed. The participant missing abc22 received a score based on seven items. Cronbach’s alpha and the mean inter-item Spearman correlation were calculated among the 14 participants with complete data. Because item selection and initial phenotype evaluation occurred in the same sample, the phenotype coefficient and nominal *P* value were considered descriptive.

### Nested Validation of the Candidate-Phenotype Pipeline

Nested leave-one-subject-out validation was applied only to the exploratory ABC item-selection and candidate-phenotype pipeline, not to the prespecified clinical, ABC subscale, ABQ, or IQ models. The complete item-screening, direction-alignment, standardization, phenotype-construction, and model-fitting process was repeated independently within each of 15 folds.

In each fold, 14 participants formed the training sample and one participant was held out. Items were eligible if they had observations from at least 10 training participants and at least two unique values. Within the training sample, the same mixed-effects model described above was fitted separately for each eligible ABC item using all seven repeated ΔMEP observations per participant. The eight eligible items with the smallest training-sample Time × Item interaction P values were selected. Training-derived item identities, directions, means, and standard deviations were then applied unchanged to the held-out participant.

A mixed-effects model fitted in the training sample was used to predict the held-out participant’s ΔMEP slope across the seven post-cTBS assessments. This predicted slope was compared with that participant’s slope estimated from the seven observed ΔMEP values. Pearson and Spearman correlations and cross-validated R² summarized held-out performance. Conventional correlation P values were descriptive because they did not account for the complete item-selection process. Fold-level item selection and held-out predictions are reported in Supplementary Table 3.

## Exploratory Multimeasure Informant Analysis

Post hoc nested analyses evaluated whether adding SRS-2 Informant T-score, ABQ Motor Total, or ABQ Total improved held-out performance. Measures were standardized using training-fold parameters and combined with the fold-specific ABC phenotype using equal weighting. ABQ Motor Total and ABQ Total were not combined because the motor score contributes to ABQ Total. Candidates were compared using Pearson and Spearman correlations, cross-validated R², root-mean-square error, and mean absolute error.

### Full-Pipeline Permutation Analysis

Participant-level permutation testing evaluated the complete nested-selection pipeline. In each of 2,000 permutations, intact 58-item ABC profiles were randomly reassigned among participants, preserving within-profile correlations and missingness while breaking their relationship with the post-cTBS outcome. The complete 15-fold pipeline, from item screening through held-out prediction, was repeated for each permutation. Observed Pearson and Spearman correlations were compared with the resulting null distributions, and two-sided empirical *P* values were calculated using the plus-one correction. All 2,000 permutations were completed successfully.

### Domain-Balanced Sensitivity Analysis

To evaluate whether separate weighting of three redundant self-injury items amplified the phenotype association, standardized items were averaged within speech, motor or behavioral-disinhibition, and self-injury domains. Each domain was standardized, and the three domain scores were averaged with equal weight.

Individual domains and two-domain combinations were also evaluated. Because only one participant endorsed any self-injury item, a sensitivity analysis excluded that participant, recomputed standardization in the remaining 14 participants, and evaluated the speech-plus-motor phenotype. These analyses were post hoc, and their mixed-model P values are descriptive. Domain and baseline-covariate sensitivity results are reported in Supplementary Table 4.

## Results

### Participants

The primary cohort comprised 19 participants, all with seven post-cTBS assessments; 15 also had concurrent caregiver-rated ABC and ABQ assessments and formed the multimeasure cohort (Supplementary Figure 1). Thirteen participants were male and six were female; mean age was 20.7 years (SD 6.9; range 15–40). Six had known genetic variants or conditions. Additional demographic, psychiatric, and treatment characteristics appear in Table 1 and Supplementary Table 1.

**Table 1.** Participant characteristics, clinical measures and TMS data quality.

| Measure | Value |
| --- | --- |
| Cohort |  |
| Full MEP cohort, N | 19 |
| Common caregiver-informant ABC + ABQ cohort, N | 15 |
| Not included in concurrent caregiver rated ABC/ABQ analyses, N | 4 |
| Demographics |  |
| Age, mean (SD), years | 20.7 (6.9) |
| Age, median [range], years | 18 [15, 40] |
| Male, N (%) | 13 (68.4) |
| Female, N (%) | 6 (31.6) |
| Known genetic variant or condition, N (%) | 6 (31.6) |
| Cognitive Characteristics |  |
| WASI-II Full Scale-4 IQ, mean (SD) | 99.5 (24.5) |
| Clinical Measures: Full Cohort |  |
| BFCRS Total | 1.2 (1.4) |
| Kanner Catatonia Severity Total | 3.8 (3.7) |
| Kanner Catatonia Examination Total | 0.5 (0.8) |
| SRS-2 Informant Total Raw Score | 94.3 (32.6) |
| Clinical Measures: Common Caregiver-Informant Cohort |  |
| ABQ Motor Total | 8.1 (5.2) |
| ABQ Total | 23.0 (12.0) |
| ABC Irritability | 7.3 (6.7) |
| ABC Social Withdrawal | 9.2 (7.3) |
| ABC Stereotypic Behavior | 4.8 (3.5) |
| ABC Hyperactivity/Noncompliance | 11.8 (6.3) |
| ABC Inappropriate Speech | 4.9 (3.3) |
| ABC Total Score | 37.9 (16.9) |
| TMS Data Quality |  |
| Post-cTBS observations, N | 133 |
| Post-cTBS assessments per participant, median [range] | 7 [7, 7] |
| Baseline MEP amplitude, mean (SD), μV | 447.5 (701.7) |
| Retained pulses per post-cTBS block, mean (SD) | 9.8 (0.4) |

Values are mean (SD), median [range], or N (%). Catatonia, SRS-2, and IQ values are from the full 19-participant cohort. ABC and ABQ values are from the same 15-participant caregiver-informant cohort.

### Psychiatric and Medical History

Attention-deficit/hyperactivity disorder was present in 15 participants. Five participants had a history of major depressive disorder, and five had a history of self-injury. No participant had previously received electroconvulsive therapy or transcranial magnetic stimulation. Prior psychiatric medication classes included antidepressants, attention-deficit/hyperactivity disorder medications, sleep or anxiety medications, and antipsychotics. Additional characteristics are reported in Supplementary Table 1.

### Clinical and Behavioral Characteristics

Catatonia symptom burden was low (Table 1). Mean SRS-2 Informant Total Raw Score was 94.3 (SD 32.6), and mean Full Scale-4 IQ was 99.5 (SD 24.5). Caregiver-informant ABQ and ABC characteristics are reported in Table 1.

### MEP Data Completeness and Trial Retention

All 19 participants contributed ΔMEP values at each of the seven post-cTBS assessments, yielding 133 complete participant-by-time observations. After within-block outlier exclusion, a mean of 9.8 of 10 MEP trials per post-cTBS assessment block was retained (SD 0.4). Mean baseline MEP amplitude was 447.5 μV (SD 701.7).

### Prespecified Clinical Predictors

BFCRS Total, KCS Total, and KCE Total did not significantly moderate the post-cTBS ΔMEP time course (all P ≥ .537; Table 2). SRS-2 Informant Total Raw Score and WASI-II Full Scale-4 IQ were also nonsignificant.

**Table 2.** Clinical predictors, informant-rated models, candidate phenotype and validation analyses.

| Predictor or Analysis | N | β or Statistic | SE | 95% CI | P | Adjusted P |
| --- | --- | --- | --- | --- | --- | --- |
| Prespecified Aim 1: Catatonia Measures |  |  |  |  |  |  |
| BFCRS Total | 19 | -0.084 | 0.183 | -0.442 to +0.274 | .646 | — |
| Kanner Catatonia Severity Total | 19 | -0.047 | 0.183 | -0.405 to +0.311 | .797 | — |
| Kanner Catatonia Examination Total | 19 | -0.113 | 0.182 | -0.470 to +0.245 | .537 | — |
| Prespecified Aim 2: Social Impairment |  |  |  |  |  |  |
| SRS-2 Informant Total Raw Score | 19 | -0.108 | 0.183 | -0.466 to +0.249 | .553 | — |
| Exploratory Clinical Predictor |  |  |  |  |  |  |
| WASI-II Full Scale-4 IQ | 19 | +0.026 | 0.183 | -0.332 to +0.385 | .885 | — |
| Exploratory Informant-Rated ABC/ABQ Family |  |  |  |  |  |  |
| ABC Inappropriate Speech | 15 | +0.688 | 0.210 | +0.276 to +1.099 | .001 | .008 |
| ABC Total Score | 15 | +0.377 | 0.219 | -0.052 to +0.805 | .085 | .595 |
| ABC Hyperactivity/Noncompliance | 15 | +0.349 | 0.219 | -0.081 to +0.779 | .111 | .668 |
| ABC Stereotypic Behavior | 15 | +0.229 | 0.221 | -0.204 to +0.662 | .300 | 1.000 |
| ABQ Total | 15 | +0.128 | 0.222 | -0.307 to +0.563 | .563 | 1.000 |
| ABC Social Withdrawal | 15 | +0.090 | 0.222 | -0.346 to +0.525 | .686 | 1.000 |
| ABC Irritability | 15 | +0.064 | 0.222 | -0.371 to +0.500 | .772 | 1.000 |
| ABQ Motor Total | 15 | +0.057 | 0.222 | -0.379 to +0.492 | .799 | 1.000 |
| Candidate Behavioral Phenotype |  |  |  |  |  |  |
| Item-weighted 8-item ABC phenotype | 15 | +0.904 | 0.200 | +0.511 to +1.297 | < .001 | — |
| Nested Validation |  |  |  |  |  |  |
| Pearson r | 15 | +0.357 | — | — | .191 | — |
| Spearman ρ | 15 | +0.446 | — | — | .095 | — |
| Cross-validated R² | 15 | +0.066 | — | — | — | — |
| Full-Pipeline Permutation Inference |  |  |  |  |  |  |
| Pearson r | 2,000 | +0.357 | — | — | .360 | — |
| Spearman ρ | 2,000 | +0.446 | — | — | .261 | — |

N denotes participants for model and nested-validation rows and permutations for full-pipeline permutation rows. β represents the Time × standardized-predictor interaction. Holm correction was applied across the eight informant-rated exploratory models comprising ABQ Motor Total, ABQ Total, five ABC subscales, and ABC Total Score. Candidate-phenotype P values are descriptive. Conventional nested-correlation P values are descriptive. The full-pipeline permutation rows report the primary two-sided empirical P values.

### Informant-Rated Clinical Measures and Post-cTBS Facilitation

ABC Inappropriate Speech was the only measure that remained significant after Holm correction across the eight-model informant-rated family (β = +0.688, SE = 0.210, 95% CI +0.276 to +1.099, raw P = .001, Holm-adjusted P = .008; Table 2 and Figure 1). ABC Total Score and Hyperactivity/Noncompliance showed positive but nonsignificant associations; both ABQ measures and the remaining ABC subscales were also nonsignificant.

**Figure 1:**
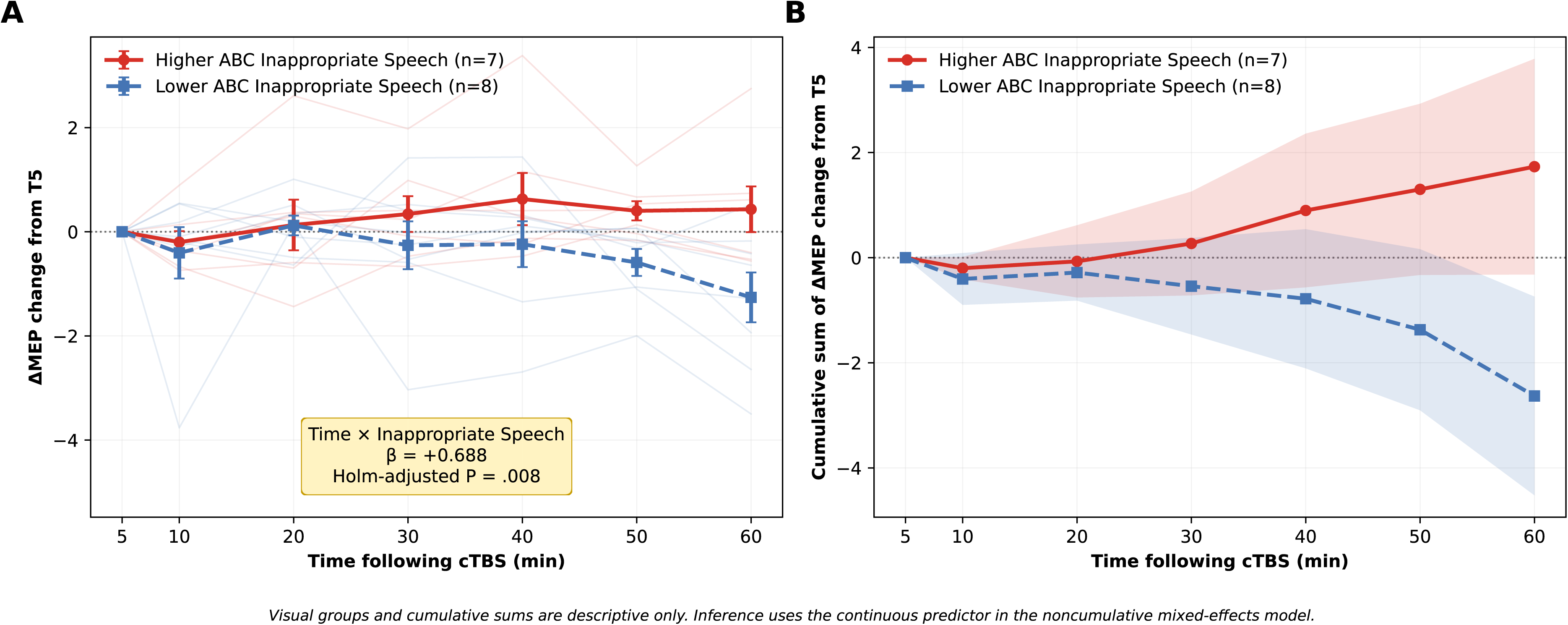
ABC Inappropriate Speech and post-cTBS motor cortical response. (A) Individual participant trajectories and group mean ± SEM for change in ΔMEP relative to the 5-min post-cTBS assessment. Participants were divided using a rank-based median split for visualization only (higher ABC Inappropriate Speech, n = 7; lower ABC Inappropriate Speech, n = 8); Study ID was used only to resolve tied scores. (B) Cumulative sum of each participant’s change from the 5-min assessment. Statistical inference used continuous, standardized ABC Inappropriate Speech scores in the noncumulative linear mixed-effects model. Higher scores were associated with progressively greater post-cTBS facilitation (Time × Inappropriate Speech interaction: β = +0.688, SE = 0.210, 95% CI +0.276 to +1.099, raw P = .001, Holm-adjusted P = .008). Error bars and shaded intervals represent SEM. The solid red line with circles indicates the higher-score group; the dashed blue line with squares and hatched uncertainty band indicates the lower-score group. ABC, Aberrant Behavior Checklist; cTBS, continuous theta-burst stimulation; MEP, motor evoked potential; SEM, standard error of the mean.

### ABC Item-Level Analysis and Candidate Behavioral Phenotype

Exploratory item-level models were fitted for all 58 ABC items. Two items remained significant after Benjamini-Hochberg correction: abc9, “Talks excessively” (β = +0.796, SE = 0.206, raw P < .001, q = .006), and abc33, “Talks to self loudly” (β = +0.686, SE = 0.210, raw P = .001, q = .032). No other item survived false-discovery-rate correction. Full item-level results are reported in Supplementary Table 2. The eight-item phenotype comprised three speech, two motor or behavioral-disinhibition, and three self-injury items (Table 3).

**Table 3.**
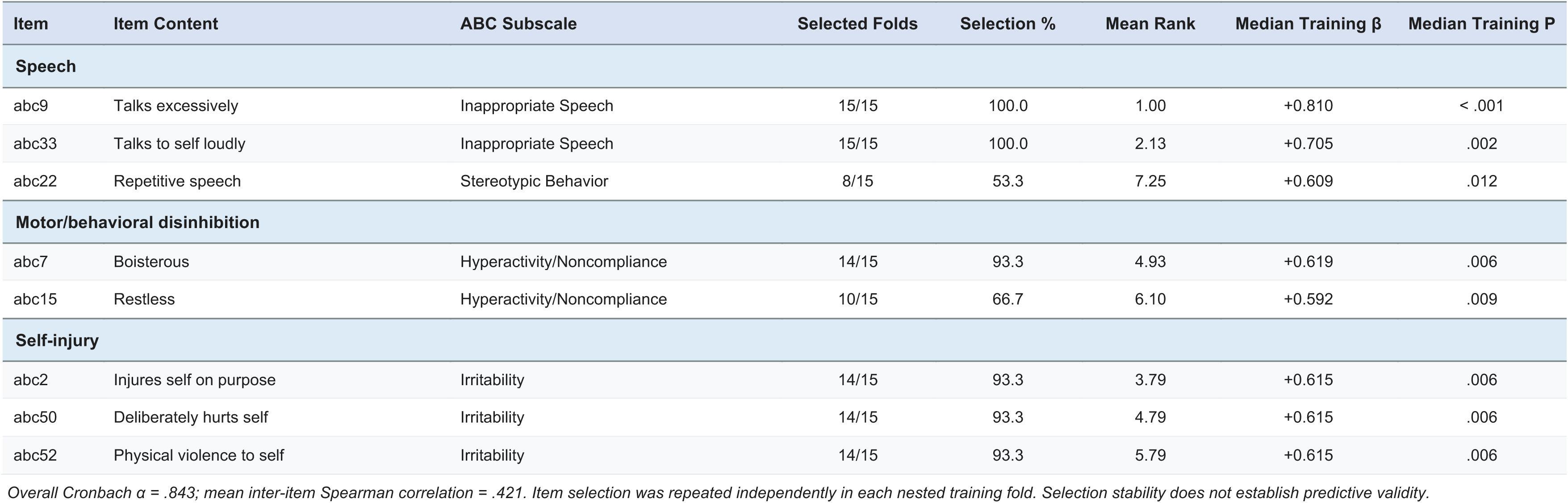
Candidate behavioral phenotype composition and nested-selection stability.

The direction-aligned eight-item candidate phenotype had a Cronbach’s α of .843 among the 14 participants with complete item data; the mean inter-item Spearman correlation was .421. The standardized Time × Candidate Phenotype interaction was large and positive (β = +0.904, SE = 0.200, 95% CI +0.511 to +1.297, descriptive P < .001; Figure 2). Because the component items were selected and evaluated in the same sample, this coefficient and its nominal P value are descriptive.

**Figure 2:**
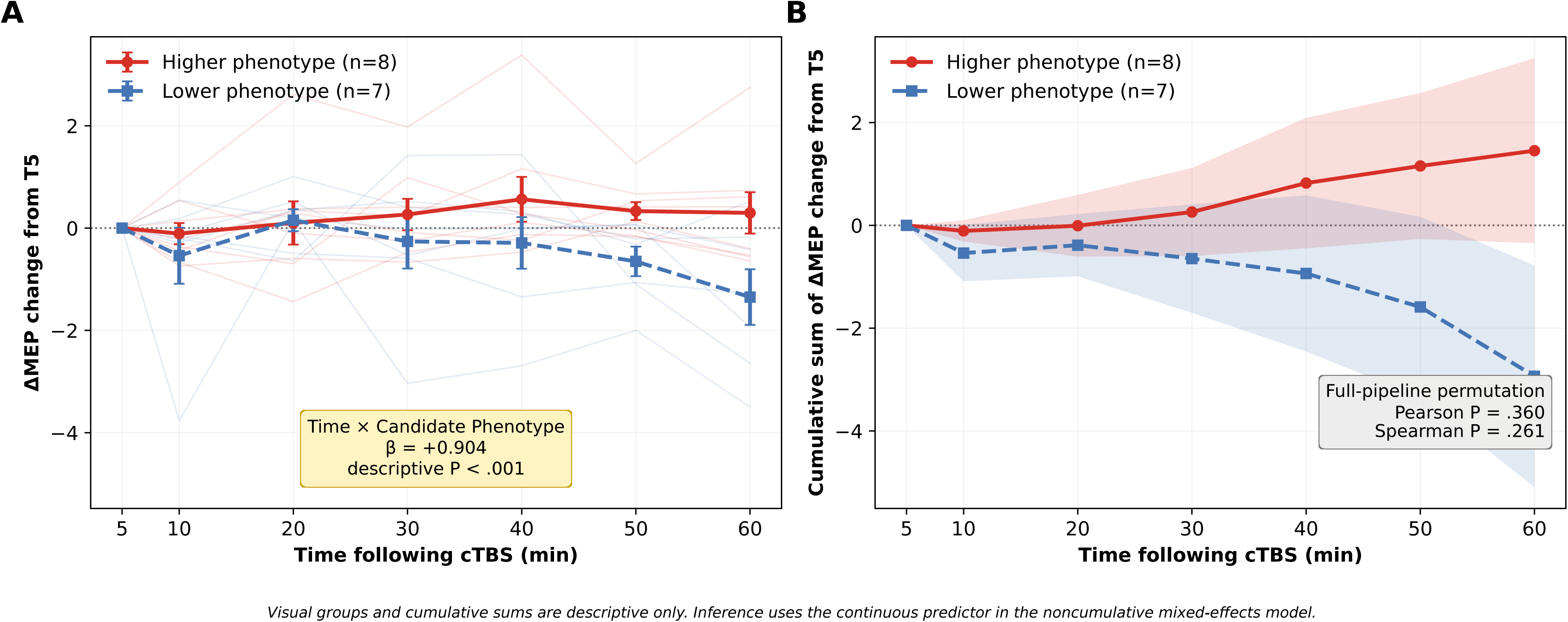
Candidate behavioral phenotype and post-cTBS motor cortical response. (A) Individual participant trajectories and group mean ± SEM for change in ΔMEP relative to the 5-min post-cTBS assessment. Participants were divided using a rank-based median split for visualization only (higher phenotype score, n = 8; lower phenotype score, n = 7); Study ID was used only to resolve tied scores. (B) Cumulative sum of each participant’s change from the 5-min assessment. Statistical inference used the continuous, standardized phenotype in the noncumulative linear mixed-effects model. The descriptive in-sample Time × Phenotype interaction was β = +0.904, SE = 0.200, 95% CI +0.511 to +1.297, descriptive P < .001. Because component items were selected and evaluated in the same sample, this mixed-model P value is descriptive. Nested held-out performance was directionally positive but nonsignificant in full-pipeline permutation testing (Pearson r = .357, two-sided empirical P = .360; Spearman ρ = .446, two-sided empirical P = .261; cross-validated R² = .066). Error bars and shaded intervals represent SEM. The solid red line with circles indicates the higher-score group; the dashed blue line with squares and hatched uncertainty band indicates the lower-score group. The candidate phenotype is hypothesis-generating. ABC, Aberrant Behavior Checklist; cTBS, continuous theta-burst stimulation; MEP, motor evoked potential; SEM, standard error of the mean.

### Nested Validation and Full-Pipeline Permutation

The nested leave-one-subject-out procedure generated held-out predictions for all 15 caregiver-informant participants. Held-out performance was directionally positive (Pearson r = .357, conventional P = .191; Spearman ρ = .446, conventional P = .095), with a cross-validated R² = .066.

The ABC-only phenotype performed best among 11 candidates; all alternatives had negative cross-validated R² values (Supplementary Table 5). abc9 and abc33 were selected in all training folds, while remaining candidate items were selected in 8–14 folds with consistent positive directions (Table 3).

All 2,000 permutations produced complete results. Neither primary two-sided empirical test was significant (Pearson P = .360; Spearman P = .261); therefore, the directionally positive held-out signal did not establish predictive validity.

## Domain-Balanced and Self-Injury Sensitivity Analysis

The equal-weighted three-domain phenotype produced an estimate nearly identical to that of the original item-weighted phenotype (β = +0.907, SE = 0.200, 95% CI +0.515 to +1.300, descriptive P < .001). A phenotype restricted to the speech and motor or behavioral-disinhibition domains also remained positively associated with the post-cTBS time course (β = +0.793, SE = 0.206, 95% CI +0.390 to +1.196, descriptive P < .001).

The three self-injury items were endorsed by the same single participant and had identical participant rankings. Their selection therefore reflected one sparse participant-level pattern rather than three independent self-injury signals. After excluding this participant and recomputing item and domain standardization in the remaining 14 participants, the speech-plus-motor phenotype remained positively associated with the post-cTBS time course (β = +0.714, SE = 0.201, 95% CI +0.320 to +1.107, descriptive P < .001).

These sensitivity analyses indicate that the broader in-sample phenotype association was neither created by assigning separate weight to three redundant self-injury items nor solely attributable to the participant endorsing self-injury. However, these analyses were post hoc and do not supersede the nonsignificant full-pipeline permutation findings.

### Additional Sensitivity Analysis

Models using log-transformed rather than raw baseline motor evoked potential amplitude produced substantively unchanged estimates. The baseline-covariate sensitivity results are reported in Supplementary Table 4.

## Discussion

Caregiver-rated inappropriate speech was associated with progressively greater post-cTBS MEP facilitation, whereas the prespecified catatonia and social-impairment hypotheses were not supported. An exploratory multi-item phenotype showed a large in-sample association but did not achieve significance in full-pipeline permutation testing. Catatonia severity was low, likely producing a floor effect, and the protocol’s structural MRI, neuronavigation, and active-motor-threshold requirements may have selected for participants with greater procedural tolerance. The null catatonia findings should therefore not be interpreted as evidence against an association with cortical plasticity; rather, they highlight the need for more inclusive neurophysiology protocols. These limitations are especially important as the field of autism research broadly has recognized that autistic individuals with intellectual disability and high support needs are systematically excluded from clinical research [10,11]. For TMS biomarker research in autism specifically, this challenge is not simply a recruitment issue; exclusion of autistic individuals across the full spectrum shapes what neurobiological conclusions can be drawn. If TMS biomarker protocols do not directly address methodological challenges of inclusion, the literature may disproportionately characterize autistic individuals with lower support needs, fewer behavioral symptoms, and lower psychiatric comorbidity, rather than the full autism spectrum.

ABC Inappropriate Speech was the only informant-rated measure to remain significant after Holm correction across the eight-model ABC/ABQ family, with higher scores associated with progressively greater post-cTBS MEP facilitation. Prior autism TMS biomarker studies have largely emphasized group differences and selected neurophysiological endpoints, often in participants without intellectual disability [12,13,15,17–23,35,55]. In contrast, the present study evaluated multiple caregiver-rated behavioral dimensions in a clinically heterogeneous sample and applied familywise correction across the eight informant-rated exploratory models. ABC Hyperactivity/Noncompliance and ABC Total Score showed positive but nonsignificant associations and did not survive correction across the eight-model informant-rated family. The ABC Inappropriate Speech finding nevertheless remains preliminary because it arose from an exploratory predictor family in a small caregiver-informant sample.

ABC Inappropriate Speech captures observable behaviors such as excessive, repetitive, or atypical verbal output, but it is not a direct measure of language ability, pragmatic communication, or speech-circuit physiology. The absence of an association in the prespecified SRS-2 Informant Total Raw Score model suggests that the observed relationship may be more closely tied to a behaviorally expressed verbal-symptom dimension than to global social impairment. This interpretation remains tentative and should be evaluated using dedicated language and communication measures.

Because the motor cortex is modulated by distributed nonmotor systems, the association with excessive, repetitive, and perseverative verbal behavior could reflect broader network-level differences in motor and behavioral regulation rather than a specific abnormality of language circuitry [56]. This framework may also be relevant to catatonia, a psychomotor syndrome involving disturbances of movement, behavior, volition, and speech, including hyperactive or excited presentations [38,41]. However, none of the prespecified catatonia measures moderated the post-cTBS response, and low catatonia severity limited the present analysis. Studies specifically recruiting autistic individuals with clinically significant catatonia are needed to determine whether catatonic symptoms are associated with altered M1 plasticity. These interpretations remain speculative and hypothesis-generating.

The exploratory item-level analysis identified an eight-item candidate phenotype spanning speech, motor or behavioral-disinhibition, and self-injury content. The phenotype showed a large in-sample association and directionally positive held-out performance, but the full-pipeline permutation tests were nonsignificant. Specifically, the nested Pearson correlation was r = .357, with a two-sided empirical P = .360, and the nested Spearman correlation was ρ = .446, with a two-sided empirical P = .261. Domain-balanced weighting and exclusion of the sole participant endorsing the self-injury items did not materially reduce the in-sample association, suggesting that redundant self-injury weighting did not create the descriptive effect. Nevertheless, because the complete selection-adjusted tests were nonsignificant, the multi-item candidate phenotype should be considered hypothesis-generating and requires evaluation in an independent sample.

Exploratory multi-measure analyses further supported the relative specificity of the ABC-derived signal. Adding SRS-2 Informant, ABQ Motor Total, or ABQ Total did not improve held-out prediction and generally reduced performance relative to the ABC-only phenotype. Neither ABQ Motor Total nor ABQ Total significantly moderated the post-cTBS trajectory in the common informant cohort. The signal therefore appeared more specific to the selected ABC behaviors, although these post hoc analyses do not establish the superiority of ABC-derived measures.

The findings may also inform future therapeutic neuromodulation studies. A recent randomized trial reported improved social communication and language outcomes after accelerated left-M1 cTBS in autistic children, using resting motor threshold and M1 targeting without complex neuronavigation [57]. These design features may reduce procedural barriers encountered in the present study. Future trials could combine baseline M1 plasticity assessment with therapeutic cTBS to evaluate whether caregiver-rated verbal-behavior symptoms or cortical-plasticity measures identify treatment-responsive subgroups. The present observational study does not establish that its plasticity measure predicts therapeutic response.

The study does not establish that inappropriate-speech symptoms cause altered M1 plasticity, that altered M1 plasticity causes speech symptoms, or that either is mechanistically linked to catatonia. ABC Inappropriate Speech is an observational caregiver-rated measure rather than a direct assessment of language ability or language circuitry. Nevertheless, caregiver-observable behavioral dimensions may be useful for biomarker development in individuals unable to complete self-report or complex task-based assessments. To our knowledge, this is the first autism TMS-derived MEP plasticity biomarker study to link post-TBS cortical modulation to a specific caregiver-rated behavioral symptom domain.

Several limitations warrant emphasis. The sample was small, with only 15 participants in the caregiver-informant analyses, limiting power, precision, and generalizability. Catatonia severity was low, and the MRI, neuronavigation, and active-motor-threshold requirements may have underrepresented participants with more severe behavioral symptoms. The observational design precludes causal inference. The candidate phenotype was selected and initially evaluated in the same sample, no external validation cohort was available, and its full-pipeline permutation tests were nonsignificant. Three self-injury items were endorsed by the same participant and did not represent independent self-injury signals, although exclusion and domain-weighting analyses did not eliminate the descriptive association. Conventional nested-correlation P values were not selection-adjusted and remain descriptive. Finally, M1 MEP modulation does not establish a mechanism involving language circuitry.

In summary, ABC Inappropriate Speech was associated with progressively greater post-cTBS MEP facilitation and was the only informant-rated measure to remain significant after Holm correction across the eight-model ABC/ABQ family. An exploratory eight-item candidate phenotype showed a large descriptive in-sample association and directionally positive held-out performance but did not reach significance in full-pipeline permutation testing and therefore remains hypothesis-generating. The prespecified catatonia and social-impairment hypotheses were not supported. Interpretation of the catatonia findings was limited by low symptom burden and the small pilot sample. These findings support independent evaluation of caregiver-rated inappropriate speech as a potential behavioral correlate of M1 plasticity and highlight the need for more inclusive biomarker protocols with reduced MRI and command-following demands. These recruitment challenges support developing inclusive TMS protocols that use resting motor threshold and reduce reliance on MRI-guided neuronavigation. In light of emerging randomized evidence that resting-threshold, M1-targeted accelerated cTBS can improve social communication and language outcomes in autistic children [57], these findings support a future precision-neuromodulation framework in which M1 cortical plasticity is tested as a biomarker for communication-related treatment response across the full autism spectrum.

## Author Contributions

Conceptualization: JRS, CC, GJ, PC. Methodology: JRS, BL, HW, JL. Investigation: JRS, MB, SM, RA, BL, SL, IB. Psychological assessment: BL. Data curation: MB, SM, RA, SL, IB. Formal analysis: JRS, HW, JL. Validation: JRS, HW, JL. Interpretation: all authors. Funding acquisition: JRS. Supervision: JRS, CC, GJ, PC. Visualization: JRS. Writing, original draft: JRS. Writing, review and editing: all authors. All authors approved the final version and agree to be accountable for the accuracy and integrity of the work.

## Data Availability

The de-identified data generated and analyzed during this study are available from the corresponding author on reasonable request, subject to institutional review and applicable participant-privacy protections.

