## Supplemental Table 1 for "Caregiver-Rated Inappropriate Speech and Post-cTBS Motor Cortical Facilitation in Autism: A Pilot Biomarker Study"

**Supplementary Table 1. Psychiatric, developmental, and treatment history of the full cohort**

| **Variable** | **N** | **%** |
| --- | --- | --- |
| **Neurodevelopmental diagnoses** | | |
| Autism spectrum disorder | 19 | 100.0 |
| DSM-5 Support Level 1 | 8 | 42.1 |
| DSM-5 Support Level 2 | 1 | 5.3 |
| DSM-5 Support Level not documented | 10 | 52.6 |
| Prior Asperger syndrome diagnosis | 3 | 15.8 |
| Prior PDD-NOS diagnosis | 2 | 10.5 |
| Intellectual disability | 1 | 5.3 |
| **Prior psychiatric history** | | |
| Prior psychiatric medication provider | 18 | 94.7 |
| Prior psychotherapy | 17 | 89.5 |
| Prior inpatient psychiatric hospitalization | 2 | 10.5 |
| Prior residential psychiatric treatment | 1 | 5.3 |
| History of self-injury | 5 | 26.3 |
| History of suicide attempts | 0 | 0.0 |
| History of trauma or abuse | 5 | 26.3 |
| History of drug or alcohol use | 3 | 15.8 |
| Family history of psychiatric diagnoses | 14 | 73.7 |
| History of state or federal agency involvement | 4 | 21.1 |
| Prior ECT or TMS | 0 | 0.0 |
| **Previous psychiatric diagnoses** | | |
| Attention-deficit/hyperactivity disorder | 15 | 78.9 |
| Major depressive disorder | 5 | 26.3 |
| Tourette syndrome | 2 | 10.5 |
| Bipolar disorder | 1 | 5.3 |
| Schizophrenia | 1 | 5.3 |
| Impulse control disorder | 1 | 5.3 |
| Other psychiatric diagnosis | 10 | 52.6 |
| Generalized anxiety disorder | 5 | 26.3 |
| Unspecified anxiety disorder | 3 | 15.8 |
| Obsessive-compulsive disorder | 2 | 10.5 |
| Conduct disorder | 1 | 5.3 |
| Catatonia, historical prior diagnosis | 1 | 5.3 |
| Psychogenic non-epileptic spells | 1 | 5.3 |
| Psychotic symptoms in the context of autism/OCD, not primary psychosis | 1 | 5.3 |
| **Prior psychiatric medication classes** | | |
| Antidepressants | 11 | 57.9 |
| ADHD medications | 11 | 57.9 |
| Sleep or anxiety medications | 9 | 47.4 |
| Antipsychotics | 4 | 21.1 |
| Mood stabilizers | 2 | 10.5 |
| Cognition-enhancing medications | 1 | 5.3 |
| Benzodiazepines | 1 | 5.3 |

*Note. Values are N and percentage of the full cohort (N = 19). Diagnoses and medication classes are not mutually exclusive. DSM-5 support levels reflect documentation at the time of diagnosis and may predate current clinical reassessment. The 'Other psychiatric diagnosis' category indicates the number of participants with at least one diagnosis listed in the indented rows and should not be summed across diagnoses. History of drug or alcohol use indicates any documented prior use and does not necessarily indicate a substance use disorder. Prior psychiatric medication classes reflect any documented historical use before study enrollment. ASD, autism spectrum disorder; DSM-5, Diagnostic and Statistical Manual of Mental Disorders, Fifth Edition; PDD-NOS, pervasive developmental disorder not otherwise specified; ADHD, attention-deficit/hyperactivity disorder; OCD, obsessive-compulsive disorder; ECT, electroconvulsive therapy; TMS, transcranial magnetic stimulation.*
