## Supplemental Table 2 for "Caregiver-Rated Inappropriate Speech and Post-cTBS Motor Cortical Facilitation in Autism: A Pilot Biomarker Study"

**Supplementary Table 2. ABC item-level analysis and candidate-phenotype psychometric characteristics**

**Panel A. Full 58-item ABC scan**

| **Rank** | **Item** | **N** | **β** | **SE** | **95% CI** | **Raw P** | **BH q** |
| --- | --- | --- | --- | --- | --- | --- | --- |
| 1 | abc9 | 15 | +0.796 | 0.206 | +0.394 to +1.199 | < .001 | .006 |
| 2 | abc33 | 15 | +0.686 | 0.210 | +0.274 to +1.097 | .001 | .032 |
| 3 | abc7 | 15 | +0.598 | 0.213 | +0.181 to +1.016 | .005 | .053 |
| 4 | abc2 | 15 | +0.592 | 0.213 | +0.174 to +1.010 | .005 | .053 |
| 5 | abc50 | 15 | +0.592 | 0.213 | +0.174 to +1.010 | .005 | .053 |
| 6 | abc52 | 15 | +0.592 | 0.213 | +0.174 to +1.010 | .005 | .053 |
| 7 | abc15 | 15 | +0.570 | 0.214 | +0.151 to +0.989 | .008 | .064 |
| 8 | abc22 | 14 | +0.579 | 0.229 | +0.130 to +1.027 | .011 | .083 |
| 9 | abc21 | 15 | +0.531 | 0.215 | +0.109 to +0.952 | .014 | .088 |
| 10 | abc26 | 15 | +0.521 | 0.215 | +0.099 to +0.943 | .016 | .090 |
| 11 | abc41 | 15 | -0.510 | 0.216 | -0.932 to -0.087 | .018 | .095 |
| 12 | abc56 | 15 | +0.465 | 0.217 | +0.040 to +0.890 | .032 | .155 |
| 13 | abc3 | 15 | +0.447 | 0.217 | +0.022 to +0.873 | .039 | .176 |
| 14 | abc17 | 15 | +0.366 | 0.219 | -0.063 to +0.795 | .094 | .391 |
| 15 | abc46 | 15 | +0.313 | 0.220 | -0.118 to +0.744 | .155 | .575 |
| 16 | abc39 | 15 | +0.308 | 0.220 | -0.123 to +0.739 | .162 | .575 |
| 17 | abc48 | 15 | -0.303 | 0.220 | -0.734 to +0.128 | .168 | .575 |
| 18 | abc1 | 15 | +0.289 | 0.220 | -0.143 to +0.720 | .190 | .611 |
| 19 | abc45 | 15 | +0.268 | 0.221 | -0.165 to +0.700 | .225 | .687 |
| 20 | abc43 | 15 | +0.235 | 0.221 | -0.198 to +0.668 | .288 | .810 |
| 21 | abc55 | 15 | +0.223 | 0.221 | -0.210 to +0.656 | .313 | .810 |
| 22 | abc8 | 15 | +0.223 | 0.221 | -0.211 to +0.656 | .314 | .810 |
| 23 | abc11 | 15 | +0.219 | 0.221 | -0.214 to +0.653 | .321 | .810 |
| 24 | abc27 | 15 | +0.200 | 0.221 | -0.233 to +0.634 | .365 | .863 |
| 25 | abc5 | 15 | +0.198 | 0.221 | -0.236 to +0.631 | .372 | .863 |
| 26 | abc10 | 15 | +0.171 | 0.222 | -0.264 to +0.605 | .441 | .947 |
| 27 | abc53 | 15 | -0.170 | 0.222 | -0.604 to +0.265 | .444 | .947 |
| 28 | abc14 | 15 | -0.165 | 0.222 | -0.599 to +0.270 | .457 | .947 |
| 29 | abc24 | 15 | +0.147 | 0.222 | -0.287 to +0.582 | .507 | .994 |
| 30 | abc13 | 15 | +0.134 | 0.222 | -0.301 to +0.568 | .547 | .994 |
| 31 | abc47 | 15 | -0.118 | 0.222 | -0.553 to +0.317 | .594 | .994 |
| 32 | abc4 | 15 | +0.115 | 0.222 | -0.320 to +0.551 | .603 | .994 |
| 33 | abc25 | 15 | -0.103 | 0.222 | -0.538 to +0.332 | .642 | .994 |
| 34 | abc54 | 15 | +0.095 | 0.222 | -0.340 to +0.530 | .669 | .994 |
| 35 | abc31 | 15 | +0.095 | 0.222 | -0.341 to +0.530 | .670 | .994 |
| 36 | abc23 | 15 | -0.078 | 0.222 | -0.513 to +0.358 | .726 | .994 |
| 37 | abc36 | 15 | +0.072 | 0.222 | -0.364 to +0.507 | .747 | .994 |
| 38 | abc16 | 15 | -0.071 | 0.222 | -0.507 to +0.364 | .749 | .994 |
| 39 | abc28 | 15 | +0.066 | 0.222 | -0.369 to +0.502 | .765 | .994 |
| 40 | abc18 | 15 | -0.060 | 0.222 | -0.496 to +0.376 | .787 | .994 |
| 41 | abc32 | 15 | -0.054 | 0.222 | -0.489 to +0.382 | .809 | .994 |
| 42 | abc51 | 15 | +0.052 | 0.222 | -0.384 to +0.488 | .815 | .994 |
| 43 | abc38 | 15 | -0.044 | 0.222 | -0.480 to +0.392 | .843 | .994 |
| 44 | abc42 | 15 | -0.038 | 0.222 | -0.473 to +0.398 | .865 | .994 |
| 45 | abc6 | 15 | +0.036 | 0.222 | -0.400 to +0.471 | .873 | .994 |
| 46 | abc30 | 15 | -0.034 | 0.222 | -0.469 to +0.402 | .880 | .994 |
| 47 | abc35 | 15 | -0.029 | 0.222 | -0.465 to +0.407 | .897 | .994 |
| 48 | abc34 | 15 | -0.025 | 0.222 | -0.461 to +0.411 | .911 | .994 |
| 49 | abc49 | 15 | -0.024 | 0.222 | -0.459 to +0.412 | .916 | .994 |
| 50 | abc44 | 15 | -0.018 | 0.222 | -0.454 to +0.418 | .935 | .994 |
| 51 | abc58 | 15 | +0.018 | 0.222 | -0.418 to +0.454 | .936 | .994 |
| 52 | abc12 | 15 | +0.015 | 0.222 | -0.421 to +0.450 | .947 | .994 |
| 53 | abc29 | 15 | -0.014 | 0.222 | -0.450 to +0.422 | .949 | .994 |
| 54 | abc57 | 15 | +0.012 | 0.222 | -0.424 to +0.448 | .957 | .994 |
| 55 | abc20 | 15 | -0.008 | 0.222 | -0.444 to +0.428 | .970 | .994 |
| 56 | abc40 | 15 | +0.007 | 0.222 | -0.429 to +0.443 | .975 | .994 |
| 57 | abc37 | 15 | +0.007 | 0.222 | -0.429 to +0.442 | .977 | .994 |
| 58 | abc19 | 15 | -0.001 | 0.222 | -0.437 to +0.435 | .997 | .997 |

*β is the standardized Time × Item interaction. Benjamini-Hochberg q values are reported across all 58 tests. FDR-significant rows are shaded.*

**Panel B. Candidate-phenotype psychometric and nested-selection characteristics**

| **Item** | **Item Content** | **Selected Folds** | **Selection %** |
| --- | --- | --- | --- |
| **Speech** | | | |
| abc9 | Talks excessively | 15/15 | 100.0 |
| abc33 | Talks to self loudly | 15/15 | 100.0 |
| abc22 | Repetitive speech | 8/15 | 53.3 |
| **Motor/behavioral disinhibition** | | | |
| abc7 | Boisterous | 14/15 | 93.3 |
| abc15 | Restless | 10/15 | 66.7 |
| **Self-injury** | | | |
| abc2 | Injures self on purpose | 14/15 | 93.3 |
| abc50 | Deliberately hurts self | 14/15 | 93.3 |
| abc52 | Physical violence to self | 14/15 | 93.3 |

*Overall Cronbach α = .843 among 14 complete cases; mean inter-item Spearman correlation = .421. Phenotype scores were available for all 15 participants under the requirement of at least six available items.*
