## Supplemental Table 3 for "Caregiver-Rated Inappropriate Speech and Post-cTBS Motor Cortical Facilitation in Autism: A Pilot Biomarker Study"

**Supplementary Table 3. Nested validation and full-pipeline permutation analysis**

**Panel A. Held-out participant-level predictions**

| **Held-out participant** | **Predicted slope** | **Observed slope** | **Prediction error** | **Selected items** |
| --- | --- | --- | --- | --- |
| 1 | -0.4836 | -0.6682 | +0.1846 | abc9, abc33, abc7, abc2, abc50, abc52, abc15, abc22 |
| 2 | +0.4808 | +0.2114 | +0.2694 | abc9, abc33, abc2, abc50, abc52, abc7, abc15, abc26 |
| 6 | -0.8741 | -0.4707 | -0.4033 | abc9, abc33, abc7, abc2, abc50, abc52, abc15, abc22 |
| 7 | +0.0668 | -0.0055 | +0.0723 | abc9, abc33, abc7, abc2, abc50, abc52, abc15, abc22 |
| 8 | -1.1308 | +0.3044 | -1.4353 | abc9, abc33, abc7, abc15, abc21, abc2, abc50, abc52 |
| 9 | -0.5305 | -0.5348 | +0.0043 | abc9, abc33, abc7, abc2, abc50, abc52, abc15, abc3 |
| 10 | -0.2094 | +1.1010 | -1.3104 | abc9, abc33, abc2, abc50, abc52, abc26, abc22, abc7 |
| 14 | -0.5264 | +1.9297 | -2.4561 | abc9, abc7, abc15, abc33, abc41, abc48, abc11, abc18 |
| 16 | -0.6971 | -1.9055 | +1.2084 | abc9, abc33, abc15, abc2, abc50, abc52, abc41, abc21 |
| 18 | -0.9767 | -1.2806 | +0.3038 | abc9, abc33, abc2, abc50, abc52, abc41, abc7, abc15 |
| 19 | -0.5456 | -1.6436 | +1.0980 | abc9, abc33, abc2, abc50, abc52, abc56, abc7, abc26 |
| 20 | -0.3784 | +0.3674 | -0.7458 | abc9, abc33, abc21, abc2, abc50, abc52, abc7, abc22 |
| 21 | +0.9559 | +0.5958 | +0.3601 | abc9, abc33, abc2, abc50, abc52, abc26, abc7, abc22 |
| 23 | -0.4969 | -1.3441 | +0.8472 | abc9, abc33, abc7, abc2, abc50, abc52, abc21, abc22 |
| 26 | +0.8118 | +0.1874 | +0.6244 | abc9, abc33, abc22, abc2, abc50, abc52, abc7, abc15 |

**Panel B. Nested item-selection stability**

| **Item** | **Item Content** | **Selected Folds** | **Selection %** | **Mean Rank** | **Median Training β** | **Median Training P** |
| --- | --- | --- | --- | --- | --- | --- |
| **Speech** | | | | | | |
| abc9 | Talks excessively | 15/15 | 100.0 | 1.00 | +0.810 | < .001 |
| abc33 | Talks to self loudly | 15/15 | 100.0 | 2.13 | +0.705 | .002 |
| abc22 | Repetitive speech | 8/15 | 53.3 | 7.25 | +0.609 | .012 |
| **Motor/behavioral disinhibition** | | | | | | |
| abc7 | Boisterous | 14/15 | 93.3 | 4.93 | +0.619 | .006 |
| abc15 | Restless | 10/15 | 66.7 | 6.10 | +0.592 | .009 |
| **Self-injury** | | | | | | |
| abc2 | Injures self on purpose | 14/15 | 93.3 | 3.79 | +0.615 | .006 |
| abc50 | Deliberately hurts self | 14/15 | 93.3 | 4.79 | +0.615 | .006 |
| abc52 | Physical violence to self | 14/15 | 93.3 | 5.79 | +0.615 | .006 |

**Panel C. Nested-validation and full-pipeline permutation summary**

| **Statistic** | **Value** |
| --- | --- |
| **Nested validation** | |
| Held-out participants | 15 |
| Observed Pearson r | +0.3574 |
| Conventional Pearson P | .191 |
| Observed Spearman ρ | +0.4464 |
| Conventional Spearman P | .095 |
| Cross-validated R² | +0.066 |
| **Permutation run** | |
| Permutations requested | 2,000 |
| Valid permutations | 2,000 |
| Invalid permutations | 0 |
| **Pearson empirical inference** | |
| Two-sided empirical P | .360 |
| One-sided empirical P | .110 |
| Directional percentile | 89.1st |
| **Spearman empirical inference** | |
| Two-sided empirical P | .261 |
| One-sided empirical P | .072 |
| Directional percentile | 92.8th |
| **Reproducibility** | |
| Random seed | 20260612 |

*Conventional correlation P values are descriptive. Two-sided empirical P values are the primary phenotype-level inferential tests. One-sided empirical P values and directional percentiles are descriptive. Every valid permutation required all 15 held-out predictions.*
