## Supplemental Table 4 for "Caregiver-Rated Inappropriate Speech and Post-cTBS Motor Cortical Facilitation in Autism: A Pilot Biomarker Study"

**Supplementary Table 4. Phenotype weighting, self-injury, and model-specification sensitivity analyses**

**Panel A. Phenotype weighting, domain, and self-injury sensitivity analyses**

| **Analysis** | **N** | **β** | **SE** | **95% CI** | **Descriptive P** |
| --- | --- | --- | --- | --- | --- |
| **Individual Domains** | | | | | |
| Speech domain | 15 | +0.792 | 0.206 | +0.389 to +1.196 | < .001 |
| Motor/behavioral disinhibition domain | 15 | +0.646 | 0.211 | +0.232 to +1.060 | .002 |
| Self-injury domain | 15 | +0.592 | 0.213 | +0.174 to +1.010 | .005 |
| **Composite Phenotypes** | | | | | |
| Domain-balanced phenotype | 15 | +0.907 | 0.200 | +0.515 to +1.300 | < .001 |
| Speech + motor domains only | 15 | +0.793 | 0.206 | +0.390 to +1.196 | < .001 |
| Speech + self-injury domains only | 15 | +0.844 | 0.203 | +0.445 to +1.242 | < .001 |
| Motor + self-injury domains only | 15 | +0.870 | 0.202 | +0.474 to +1.266 | < .001 |
| **Exclusion Sensitivity** | | | | | |
| Speech + motor after excluding self-injury endorser | 14 | +0.714 | 0.201 | +0.320 to +1.107 | < .001 |

*These analyses were post hoc, and their mixed-model P values are descriptive.*

**Panel B. Baseline-covariate specification sensitivity**

| **Predictor** | **N** | **Raw-baseline β** | **Raw-baseline SE** | **Raw-baseline P** | **Log-baseline β** | **Log-baseline SE** | **Log-baseline P** |
| --- | --- | --- | --- | --- | --- | --- | --- |
| ABQ Motor Total | 15 | +0.057 | 0.222 | .799 | +0.057 | 0.222 | .799 |
| ABQ Total | 15 | +0.128 | 0.222 | .563 | +0.128 | 0.222 | .563 |
| ABC Irritability | 15 | +0.064 | 0.222 | .772 | +0.064 | 0.222 | .772 |
| ABC Social Withdrawal | 15 | +0.090 | 0.222 | .686 | +0.090 | 0.222 | .686 |
| ABC Stereotypic Behavior | 15 | +0.229 | 0.221 | .300 | +0.229 | 0.221 | .300 |
| ABC Hyperactivity/Noncompliance | 15 | +0.349 | 0.219 | .111 | +0.349 | 0.219 | .111 |
| ABC Inappropriate Speech | 15 | +0.688 | 0.210 | .001 | +0.688 | 0.210 | .001 |
| ABC Total Score | 15 | +0.377 | 0.219 | .085 | +0.377 | 0.219 | .085 |

*Primary raw-baseline and log-baseline MEP covariate specifications produced substantively unchanged estimates. Values are rounded to three decimal places.*
