## Supplemental Table 5 for "Caregiver-Rated Inappropriate Speech and Post-cTBS Motor Cortical Facilitation in Autism: A Pilot Biomarker Study"

**Supplementary Table 5. Nested held-out performance of SRS-2, ABC, and ABQ candidate combinations**

| **Rank** | **Candidate** | **Components** | **Held-out N** | **Pearson r** | **Spearman ρ** | **Cross-validated R²** | **RMSE** | **MAE** | **Δ cross-validated R² vs ABC phenotype** |
| --- | --- | --- | --- | --- | --- | --- | --- | --- | --- |
| 1 | ABC phenotype | ABC phenotype | 15 | +0.357 | +0.446 | +0.066 | 0.988 | 0.755 | +0.000 |
| 2 | ABC phenotype + SRS-2 | ABC phenotype + SRS-2 Informant T-score | 15 | +0.212 | +0.293 | -0.195 | 1.117 | 0.854 | -0.261 |
| 3 | SRS-2 + ABQ Total | SRS-2 Informant T-score + ABQ Total | 15 | -0.432 | -0.371 | -0.214 | 1.126 | 0.878 | -0.280 |
| 4 | SRS-2 + ABQ Motor | SRS-2 Informant T-score + ABQ Motor Total | 15 | -0.657 | -0.618 | -0.223 | 1.130 | 0.906 | -0.289 |
| 5 | ABC phenotype + ABQ Total | ABC phenotype + ABQ Total | 15 | +0.177 | +0.218 | -0.237 | 1.137 | 0.928 | -0.303 |
| 6 | SRS-2 Informant | SRS-2 Informant T-score | 15 | -0.550 | -0.461 | -0.244 | 1.140 | 0.875 | -0.310 |
| 7 | ABQ Total | ABQ Total | 15 | -0.687 | -0.632 | -0.265 | 1.150 | 0.940 | -0.331 |
| 8 | ABC phenotype + SRS-2 + ABQ Total | ABC phenotype + SRS-2 Informant T-score + ABQ Total | 15 | +0.127 | +0.204 | -0.267 | 1.150 | 0.894 | -0.333 |
| 9 | ABQ Motor Total | ABQ Motor Total | 15 | -0.896 | -0.925 | -0.289 | 1.160 | 0.969 | -0.355 |
| 10 | ABC phenotype + SRS-2 + ABQ Motor | ABC phenotype + SRS-2 Informant T-score + ABQ Motor Total | 15 | +0.093 | -0.004 | -0.360 | 1.192 | 0.921 | -0.426 |
| 11 | ABC phenotype + ABQ Motor | ABC phenotype + ABQ Motor Total | 15 | +0.116 | +0.057 | -0.367 | 1.195 | 0.935 | -0.433 |

*All candidates used the same 15 participants. ABC item selection and phenotype construction were repeated within each training fold. SRS-2 Informant, ABQ Motor Total, and ABQ Total were standardized using training-fold parameters. ABQ Motor Total and ABQ Total were not entered into the same composite because ABQ Motor Total contributes to ABQ Total. The candidate comparison was post hoc. Negative cross-validated R² values indicate worse squared-error performance than prediction using the mean observed held-out slope.*
