## Supplemental Table 6 for "Caregiver-Rated Inappropriate Speech and Post-cTBS Motor Cortical Facilitation in Autism: A Pilot Biomarker Study"

**Supplementary Table 6. Analysis hierarchy, analytic samples, outcomes, multiplicity procedures, and inferential status**

| **Analysis** | **Status** | **N / folds** | **Predictor(s)** | **Outcome and observations** | **Primary coefficient or metric** | **Multiplicity, validation, and interpretation** |
| --- | --- | --- | --- | --- | --- | --- |
| **Prespecified clinical aims** | | | | | | |
| Aim 1: Catatonia severity | Prespecified | 19 | BFCRS Total; KCS Total; KCE Total, in separate models | Seven repeated post-cTBS ΔMEP observations per participant (5–60 min) | Time × Predictor | Three separate prespecified models; exact two-sided P values reported. |
| Aim 2: Social impairment | Prespecified | 19 | SRS-2 Informant Total Raw Score | Seven repeated post-cTBS ΔMEP observations per participant (5–60 min) | Time × Predictor | Single prespecified model; exact two-sided P value reported. |
| **Planned exploratory clinical models** | | | | | | |
| Cognitive ability | Planned exploratory | 19 | WASI-II Full Scale-4 IQ | Seven repeated post-cTBS ΔMEP observations per participant (5–60 min) | Time × Predictor | Exploratory model reported separately from the ABC/ABQ family. |
| Informant-rated ABC and ABQ measures | Planned exploratory | 15 | ABQ Motor Total; ABQ Total; five ABC subscales; ABC Total Score | Seven repeated post-cTBS ΔMEP observations per participant (5–60 min) | Time × Predictor | Holm correction across eight caregiver-informant models. |
| **Exploratory ABC item and phenotype analyses** | | | | | | |
| ABC item-level screen | Exploratory | 15 | Each of 58 ABC items, evaluated in a separate model | Seven repeated post-cTBS ΔMEP observations per participant (5–60 min) | Time × Item | Benjamini-Hochberg correction across 58 tests; item-level screen, not factor analysis. |
| Observed-sample candidate phenotype | Post hoc exploratory | 15 | Equal-weighted score from eight ABC items with the smallest raw Time × Item P values | Seven repeated post-cTBS ΔMEP observations per participant (5–60 min) | Time × Phenotype | Coefficient and nominal P value are descriptive because selection and evaluation used the same sample. |
| **Nested validation and permutation inference** | | | | | | |
| Nested candidate-phenotype validation | Post hoc validation | 15 LOSO folds | Fold-specific eight-item ABC phenotype selected in 14 training participants | Held-out ΔMEP slope estimated from seven post-cTBS observations | Pearson r; Spearman ρ; cross-validated R² | Complete selection pipeline repeated within each fold; applied only to the candidate-phenotype pipeline. |
| Full-pipeline permutation analysis | Post hoc inferential validation | 15; 2,000 permutations | Intact 58-item ABC profiles randomly reassigned among participants | Held-out slope predictions from the complete nested pipeline | Two-sided empirical P values for Pearson and Spearman | Complete 15-fold pipeline repeated in every permutation; two-sided empirical P values are primary. |
| **Additional post hoc and sensitivity analyses** | | | | | | |
| Exploratory multimeasure informant analysis | Post hoc | 15 LOSO folds | ABC phenotype alone or combined with SRS-2, ABQ Motor Total, or ABQ Total | Held-out ΔMEP slope estimated from seven observations | r; ρ; cross-validated R²; RMSE; MAE | Training-fold standardization; candidate comparisons are exploratory. |
| Domain-balanced phenotype sensitivity | Post hoc sensitivity | 15 | Equal-weighted speech, motor/behavioral-disinhibition, and self-injury domains | Seven repeated post-cTBS ΔMEP observations per participant (5–60 min) | Time × Phenotype | Mixed-model P values are descriptive. |
| Speech-plus-motor phenotype | Post hoc sensitivity | 15 | Equal-weighted speech and motor/behavioral-disinhibition domains | Seven repeated post-cTBS ΔMEP observations per participant (5–60 min) | Time × Phenotype | Mixed-model P value is descriptive. |
| Self-injury participant exclusion | Post hoc sensitivity | 14 | Speech-plus-motor phenotype recomputed after excluding the sole self-injury endorser | Seven repeated post-cTBS ΔMEP observations per participant (5–60 min) | Time × Phenotype | Mixed-model P value is descriptive. |
| Baseline-MEP covariate sensitivity | Model-specification sensitivity | Applicable samples | Log-transformed rather than raw baseline MEP amplitude | Seven repeated post-cTBS ΔMEP observations per participant (5–60 min) | Time × Predictor | Compared with corresponding primary model specifications. |

*Note. All mixed-effects models used all seven post-cTBS ΔMEP observations per participant; no peak, average, or single post-cTBS time point was selected. All primary mixed-effects models included baseline MEP amplitude, time, the predictor, and the Time × Predictor interaction as fixed effects, with a participant-specific random intercept. Positive interaction coefficients indicate progressively greater facilitation or less suppression. Nested leave-one-subject-out validation and full-pipeline permutation testing applied only to the exploratory ABC item-selection and candidate-phenotype pipeline. ABC, Aberrant Behavior Checklist; ABQ, Attenuated Behavior Questionnaire; BFCRS, Bush-Francis Catatonia Rating Scale; ΔMEP, motor-evoked-potential log-response ratio; KCE, Kanner Catatonia Examination; KCS, Kanner Catatonia Severity; LOSO, leave-one-subject-out; MAE, mean absolute error; RMSE, root-mean-square error; SRS-2, Social Responsiveness Scale, Second Edition; WASI-II, Wechsler Abbreviated Scale of Intelligence, Second Edition.*
