## Supplemental Figure 1 for "Caregiver-Rated Inappropriate Speech and Post-cTBS Motor Cortical Facilitation in Autism: A Pilot Biomarker Study"

### Supplementary Figure 1: Participant flow through enrollment, diagnostic TMS procedures, and analytic cohorts

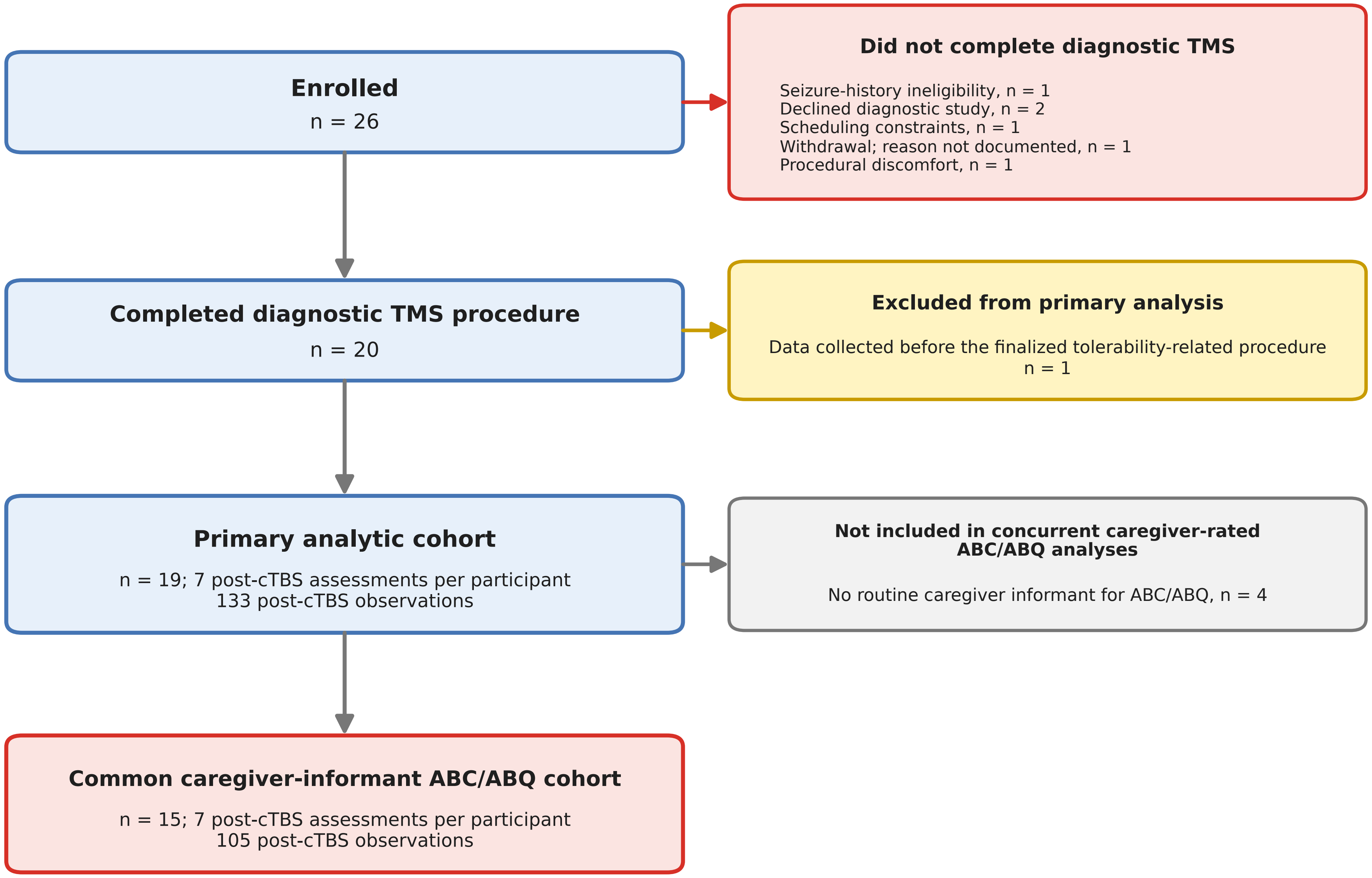


*No randomization, treatment allocation, or concurrent comparator was used. All participants completing the finalized procedure followed the same diagnostic TMS protocol.*

ABC, Aberrant Behavior Checklist; ABQ, Attenuated Behavior Questionnaire; cTBS, continuous theta-burst stimulation; TMS, transcranial magnetic stimulation.
